# The crucial role of titin in fetal development: recurrent miscarriages and bone, heart, and muscle anomalies characterize the severe end of titinopathies spectrum

**DOI:** 10.1101/2022.10.28.22281590

**Authors:** Maria Francesca Di Feo, Victoria Lillback, Manu Jokela, Meriel McEntagart, Tessa Homfray, Elisa Giorgio, Guido C Casalis Cavalchini, Alfredo Brusco, Maria Iascone, Luigina Spaccini, Patrizia D’Oria, Marco Savarese, Bjarne Udd

## Abstract

**Background:** Titin truncating variants (TTNtv) have been associated with several forms of myopathies and/or cardiomyopathies. In homozygosity or in compound heterozygosity they cause a wide spectrum of recessive phenotypes with a congenital or childhood onset. Most recessive phenotypes showing a congenital or childhood onset have been described in subjects carrying biallelic TTNtv in specific exons. However, *TTN* is not yet included in many NGS panels for congenital musculoskeletal anomalies and dysmorphisms, and karyotype or chromosomal microarray analyses are often the only tests performed when prenatal anomalies are identified. Thereby, many cases caused by *TTN* defects might be missed in the diagnostic evaluations. In this study, we aimed to dissect the most severe end of the titinopathies spectrum.

**Methods:** We performed a retrospective study analysing an international cohort of 93 published and 10 unpublished cases carrying biallelic TTNtv.

**Results:** We identified recurrent clinical features in antenatal and congenital recessive titinopathies, including fetal akinesia, arthrogryposis, facial dysmorphisms, joint, bone, and heart anomalies resembling complex, syndromic phenotypes.

**Conclusion:** We suggest *TTN* to be carefully evaluated in any diagnostic process involving patients with the mentioned signs. This step will be essential to improve diagnostic performance, expand our knowledge, and optimize prenatal genetic counseling.

**Key message:** Truncating variants in the titin gene (*TTN*) have been associated with a large spectrum of skeletal muscle diseases with or without cardiac involvement. The current work demonstrates that truncating biallelic variants in specific exons of *TTN* result in severe developmental anomalies, affecting not only the muscular systems but also bone, heart, and other organs, with a quite high rate of miscarriages and dysmorphisms. Thereby, we suggest TTN variants to be carefully evaluated in any diagnostic process involving patients with the mentioned signs.

## Introduction

The *TTN* gene (Online Mendelian Inheritance in Man database [OMIM] #188840) includes 364 exons (363 coding exons and the first non-coding exon) and codes for titin, the largest protein in our body.^1^ Titin forms the third myofilament structure spanning the sarcomere from the Z-disc to the M-band in both skeletal and cardiac muscle. The titin I-band portion works as a molecular spring, giving the muscle its elastic properties.^1,2^ Besides the mechanical properties titin also has a role as a mechanosensor serving various signalling functions.^3^

Titin transcripts have a complex splicing pattern with several known isoforms.^4^ The canonical skeletal muscle isoform N2A is reported to include 312 exons and five isoforms are reported to have a cardiac expression. The longest of these is the N2BA which contains 311 exons.^1^ The theoretical isoform that includes all 363 coding exons is the virtual TTN metatranscript (NM_001267550).^5^ Exons not contained to any larger extent in adult skeletal or cardiac muscle isoforms are referred to as metatranscript-only exons.^6^

Pathogenic *TTN* variants cause a wide range of skeletal myopathies and cardiomyopathies or combinations of both, varying by their mode of inheritance, age of onset, muscle involvement, severity, and rate of progression.^7–12^

Some genotype-phenotype correlations in titinopathies, depending on the location of the variants, have been shown, e.g. the adult-onset hereditary myopathy with early respiratory failure (HMERF) [OMIM #60369] is specifically caused by missense variants in the exon 344, and the late onset dominant distal myopathy, tibial muscular dystrophy (TMD, Udd myopathy), with heterozygous variants in the last exon 364.^13,14^

Similarly, a correlation between location of titin truncating variants (TTNtv) and the clinical manifestation of recessive titinopathies has been proposed. ^6,15^ Variants causing premature stop codons in the I- and A-band mainly result in nonsense mediated decay. This is supported by the absence of the predicted truncated protein product in skeletal muscle of most patients with these variants.^6^ On the contrary, premature stop codons in the M-band (last six exons 359-364) result in a near-full length protein that is probably able to integrate into the sarcomere. There is a clear relationship between the position of the truncation in the M-band and the phenotype in patients. Very terminal biallelic truncating variants in the last two exons 363-364 cause juvenile-early adult onset recessive distal titinopathy.^16^ On contrary, patients carrying a TTNtv at the beginning of the M-band (exon 359) in homozygosity or in compound heterozygosity with a second TTNtv in a canonical exon out of the M-band, have a congenital onset resulting in neonatal hypotonia, leading to a severe form of congenital titinopathy. ^6,17,18^ Finally, a truncating variant in a metatranscript-only exon (metaTTNtv) in homozygosity, or in compound heterozygosity with another class of truncating variant causes arthrogryposis congenita and severe axial hypotonia as a form of congenital amyoplasia. ^15,19,20^

Our study, analyzing clinical and molecular data from a cohort of novel and previously described recessive titinopathy cases with congenital anomalies or dysmorphisms, proves that biallelic titin pathogenic variants cause recognizable fetal and developmental defects. All the newly reported patients show a severe prenatal or congenital phenotype leading to fetal, perinatal, or infantile death, thus describing the most severe end of the titinopathies.

## Patients and methods

### Recruitment

We collected either cases from obstetrical and neonatology units of different international hospitals or cases that have been brought to our attention by direct request for counselling.

### Clinical Features Analysis

The cases have been clinically assessed by gynecologists experienced in prenatal diagnostics; in three cases autopsy was performed after voluntary abortion or fetal death (F4-II.1; F4-II.2; F6-II.3; F6-II.4). Cases born alive were hospitalized in neonatal intensive care units before death; case F5-II.1, who died in childhood, was followed by both pediatricians and neurologists.

### Molecular Genetic Studies

Probands’ DNA was analyzed using gene panels or exome sequencing (ES). Sequencing data was analyzed using standard bioinformatic pipelines aiming at the identification of single nucleotide variants, small insertions or deletion (indels). In all cases, segregation analysis (via Sanger or massive parallel sequencing, MPS) was performed to confirm the phase of the variants.

All the variants had been evaluated following the ACMG/AMP criteria for variants interpretation using the default settings in Varsome (version 5.6 on 14/10/2022).^21^ Variants are reported in the manuscript using the inferred meta-transcript NM_001267550.1.

### Previously reported cases

Medical literature was searched using PubMed (search terms: titin, TTN, titinopathy, congenital myopathy, muscular dystrophy; last search undertaken on 1 October 2022). We included previously reported patients with antenatal, congenital, infantile or childhood onset (< 2 years of age) recessive titinopathy carrying biallelic TTNtv classified as pathogenic or likely pathogenic according to current guidelines for whom molecular and clinical findings were available (n = 93 cases) (Supplementary Table 1).

## Results

We reported 10 cases from 6 unrelated families (Table 1) showing different TTNtv combinations. Four cases carried a heterozygous variant in exon 359 in combination with a TTNtv in a canonical exon (Z-disk, I-band or A-band); one case was a compound heterozygous for two TTNtv in exon 359, and five cases carried a TTNtv in a metatranscript-only exon in combination with a TTNtv in a canonical exon (A-band). Molecular and clinical details of these 10 cases are summarized in Table 1.

**Table 1.**
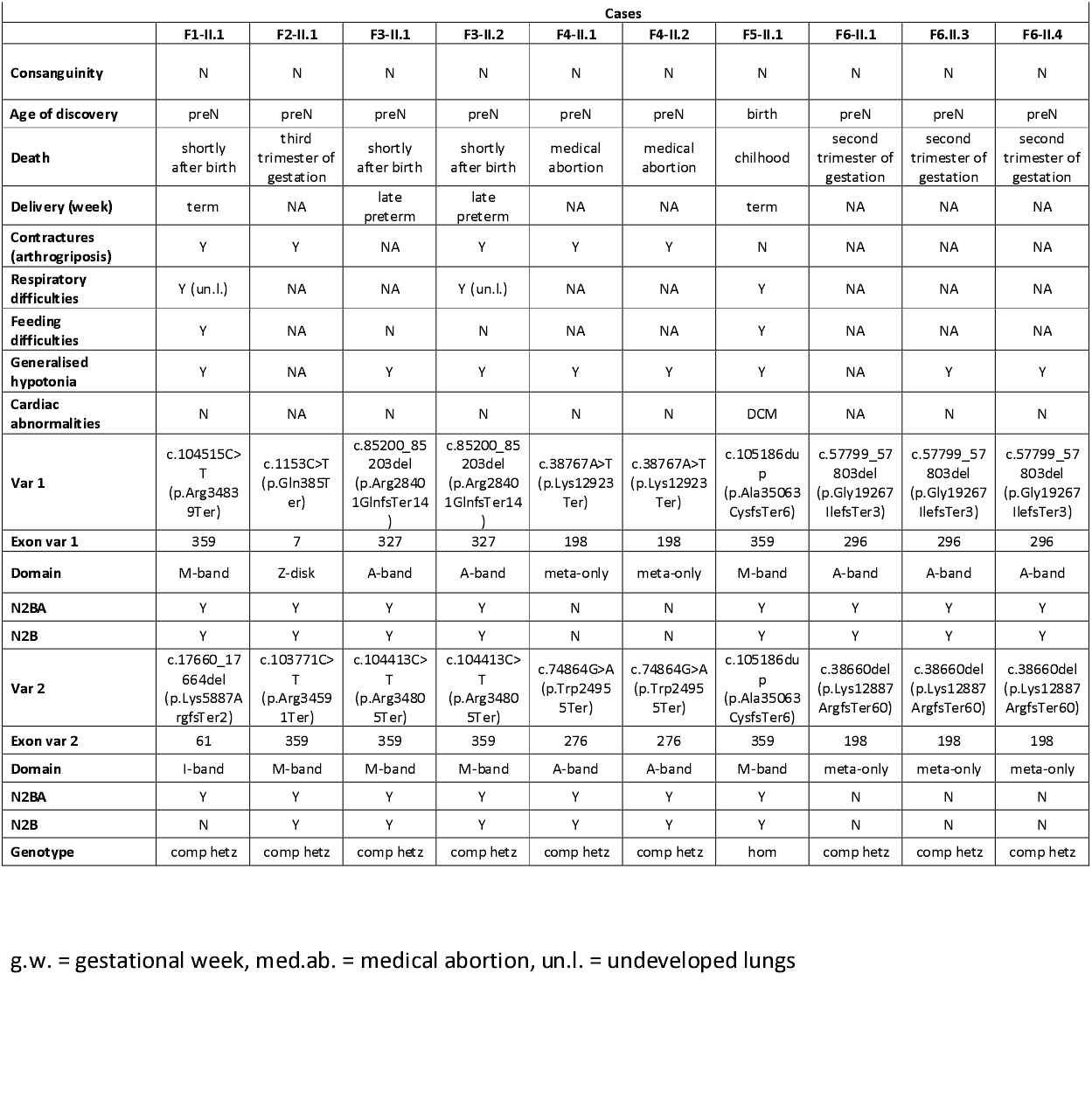
Summarized genotypes and phenotypes of the 10 unpublished cases.

### Family history and first-level analyses

Family history, as depicted by the pedigrees in Supplementary Figure 1, was negative in all the cases, except for one grandparent (family 3) who was reported to have dilated cardiomyopathy although he was not genetically investigated.

**Figure 1.**
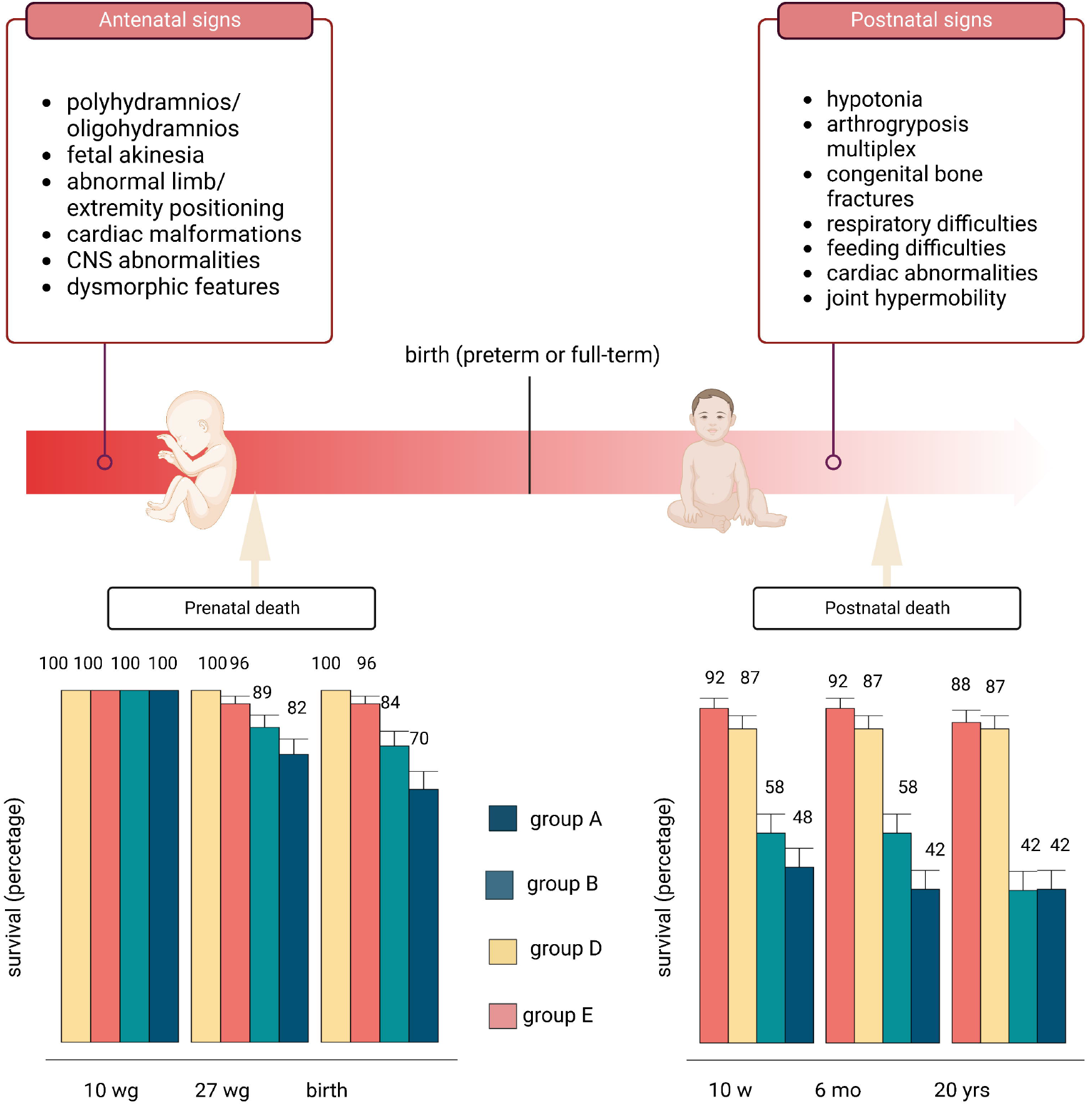
Ultrasound examination images of case F6-II.3 (first three images) and F6-II.4 (last three images). Abnormality of corpus callosum, placenta previa and limb abnormalities in case F6-II.3 (A,B,C) and severe arthrogryposis in case F6-II.4 (D,E,F) can be observed.

In 3 out of 6 families, exome sequencing of the trio was not the first-tier test. In two cases (family 3 and 6), a sequencing test covering titin was performed following the re-occurrence of the disease in a second sibling. All the other genetic analyses performed (karyotype, CGH array, in one case also *SMN1* sequencing) gave normal results. No other clinically relevant disease-causing variants were identified by trio sequencing analysis.

### Unreported cases with at least one metatranscript-only variant

All five patients carrying a TTNtv in a metatranscript-only exon died in uterus spontaneously or following a voluntary abortion procedure due to severe ultrasound-detected abnormalities (n=2), approximately between the 25 and the 35 weeks of gestation. In all cases severe signs such as fetal akinesia and limb contractures were found at ultrasound examination. In particular, in case F4-II.2, abnormal villous maturation was reported at autopsy, in addition to the typical arthrogryposis multiplex phenotype (images available upon request). In case F6-II.3, ultrasound examination in the second trimester of gestation showed abnormalities of corpus callosum, absent movements on prolonged observation, upper and lower limbs in constant flexion, immobile extremities, flattened thorax anteriorly with normal thorax/abdomen ratio. In case F6-II.4 (Figure 1), severe contractures of upper and lower limbs were found in 2 different ultrasound examinations at 16 and 20 weeks. All metatranscript-only cases showed severe muscle hypoplasia, skeletal muscle damage and rarefaction of muscle fibres at the anatomopathological examination.

### Unreported cases with an exon 359 truncating variant

Four cases from 3 unrelated families (families 1, 2, 3) with a combination of TTNtv in exon 359 and a TTNtv in a canonical exon showed severe abnormalities on ultrasound examination, including hydrops fetalis and polyhydramnios. One fetus (F2-II.1) died in uterus in the third trimester of gestation, while two others (F3-II.1, F3-II.2) were born preterm, and one was born at early-term (F1-II.1). They all presented arthrogryposis, one of them joint dislocation, and two of them showed undeveloped lungs. No congenital cardiac defect was reported in our cohort. Only the patient carrying two TTNtv in exon 359 (F5-II.1), who survived until childhood, developed dilated cardiomyopathy during infancy. All the presented cases (F1-II.1, F3-II.1, F3-II.2, F5-II.1) with at least one TTNtv in exon 359 who were born alive died of respiratory failure.

### Clinical features of antenatal and congenital titinopathies: evidence in a wider context

Ninety-three recessive cases due to biallelic TTNtv with congenital or early onset (< 2 years of age) titinopathy have been reported in the literature (Supplementary Table 1). For the purpose of this paper, we considered only those cases for which molecular and clinical findings were available. In addition, we added to the analysis the 10 new cases of recessive titinopathies described above, to refine a tentative genotype-phenotype correlation.

Of the 103 analyzed cases, sixty-three (61%) were alive at the time of data collection. For a proper analysis of the antenatal and perinatal signs and symptoms, we grouped and analysed cases according to their genotype. The overall survival rates at birth, in childhood, and in adolescence according to genotype are shown in Figure 3.

### Evidence on metaTTNtv

Thirteen cases have bi-allelic nonsense or indels causing a premature stop-codon in metatranscript-only exons. Twenty cases have a nonsense or an indel causing a frameshift in a metatranscript-only in compound heterozygosity with a nonsense or an indel variant causing a premature stop codon in a canonical exon. Overall, metaTTNtv cases (group A, Suppl Table 1) show the lowest survival rate in the prenatal and perinatal period in the entire cohort, as only fourteen out of thirty-three patients (42%) were alive 6 months post partum (Figure 3). This group also shows a consistent prevalence of reported abnormal prenatal signs such as fetal akinesia (55%, n=18), polyhydramnios (18%, n=6) and fetal hydrops (18%, n=6) (Figure 2). Micrognathia, retrognathia, facial anomalies, and other dysmorphisms have been reported in 45% (n=15) of the cases. At birth, almost all of them (85%, n=28) presented with severe contractures, especially in distal limbs, and they were described as having arthrogryposis multiplex congenita or as distal arthrogryposis. A large proportion of live infants also had generalised hypotonia (78%, n=18), respiratory difficulties requiring intubation (35%, n=8), and feeding difficulties, requiring nasogastric tube feeding (30%, n=7); some of them (n=9) died after a few weeks mainly of respiratory insufficiency. Also, 22% of the infants (n=5) reported congenital bone fractures at birth. Only one case reported unspecified cardiac anomalies at birth. Two infants who died after the delivery showed hypoplastic hearts at autopsy.

**Figure 2.**
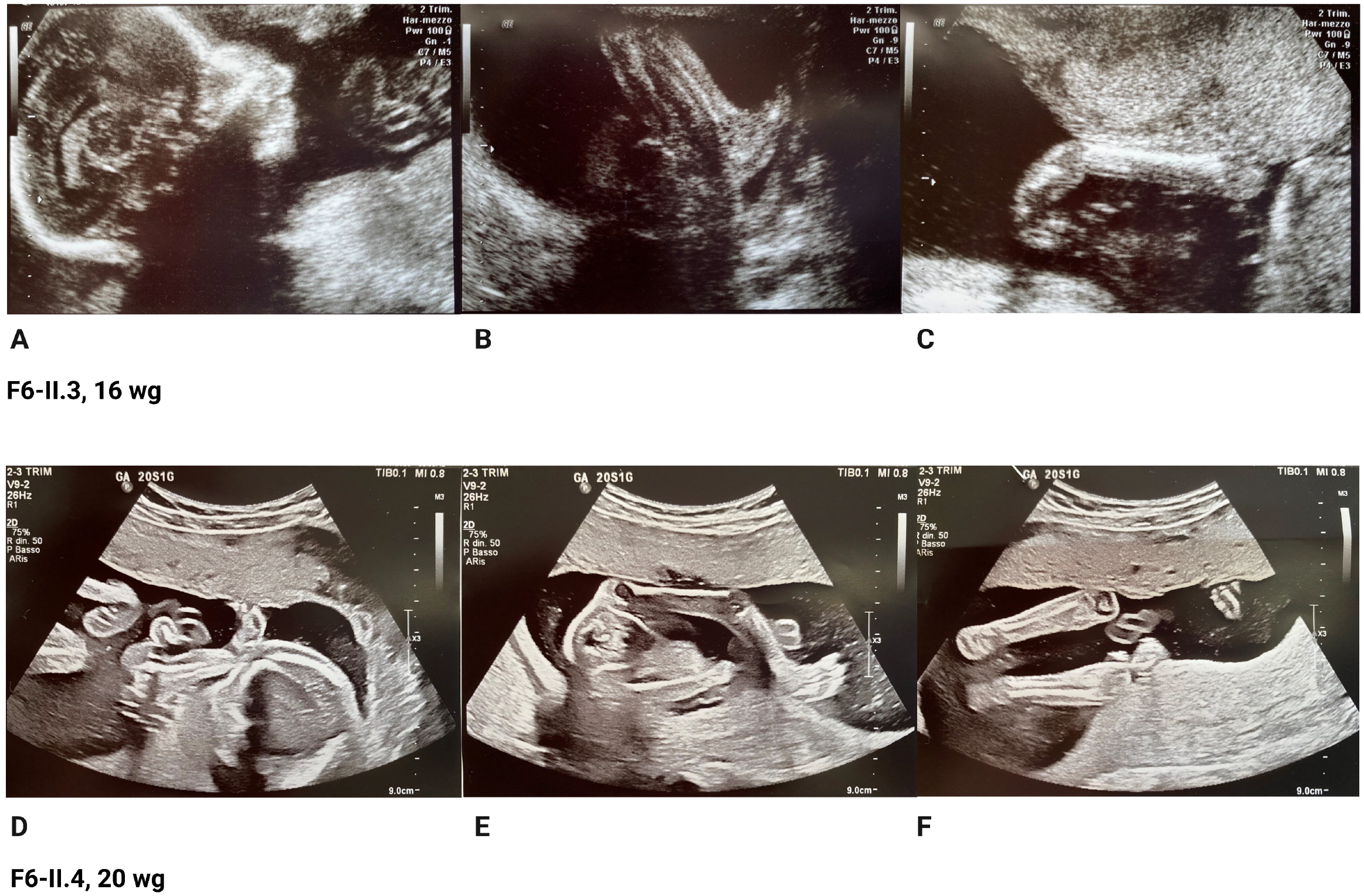
Analysis of the clinical findings according to the genotype: group A = metaTTNtv, group B = exon 359 TTNtv, group C = exon 360-363 TTNtv, group D = canonical TTNtv with splice variant, group E = metaTTNtv with splice variant. In-depth descriptions of genotypes are provided in the manuscript. Chi-square test was performed and p-values have been displayed next to the clinical signs when there is significant difference between the groups. P-value equal to or less than 0.05 = *; p-value equal to or less than 0.01 = **; p-value equal to or less than 0.001 = ***

**Figure 3.**
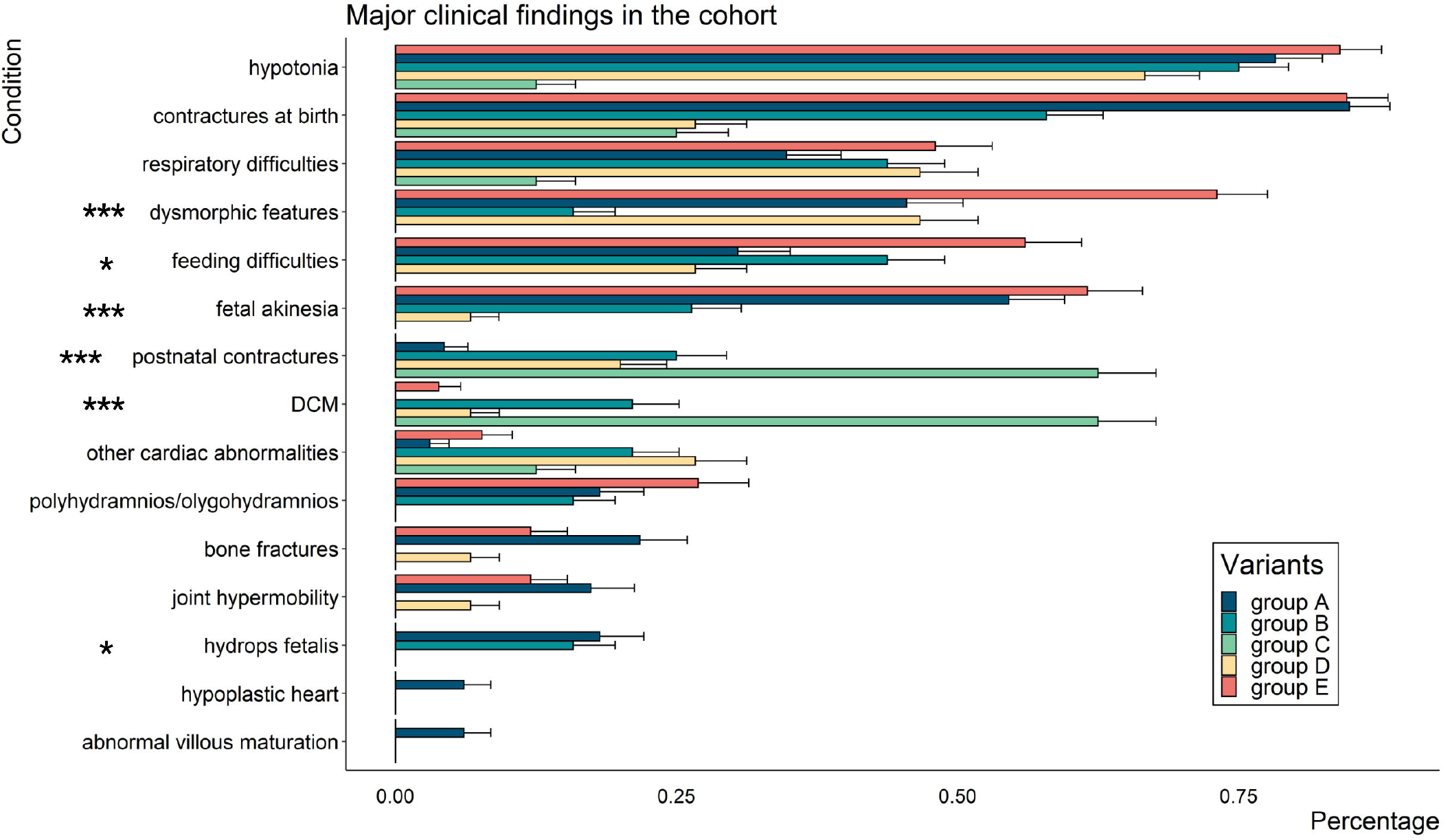
Description of the main signs suggestive of prenatal titinopathy in fetuses and infants. Survival rates in percentages are shown for the following groups: group A = metaTTNtv, group B = exon 359 TTNtv, group D = canonical TTNtv with splice variant, group E = metaTTNtv with splice variant. In-depth descriptions of genotypes are provided in the manuscript. Created with BioRender.com

### Evidence on TTNtv in exon 359

Nineteen patients carry a nonsense or an indel causing a frameshift in exon 359 in homozygosity or in compound heterozygosity with second variant in a canonical exon causing a premature stop codon out of the M-band (group B, Suppl Table 1). Of these, three cases (16%) died in uterus, two (10,5%) by medical abortion for severe clinical findings, one (5%) for miscarriage at the 35^th^ weeks. Among the survived cases, five died (31%) up to six months after birth, while other three (27% of the alive cohort) died in juvenile age (< 30 y.o.) due to DCM. Fifteen cases (79%) reported limb contractures, but in four of them, the onset was clearly postnatal.

Two cases carrying out-of-frame deletions in exon 359 in compound heterozygosity with a metaTTNtv have been described (group F, Suppl table 1). They presented with hypotonia at birth, and they were both alive at the data collection. One of them developed contractures after birth.

### Evidence on TTNtv in exons 360-363

Eight previously published cases reported out-of-frame indels in exons 360-363 (group C, Suppl Table 1). They had mostly a childhood onset (at 1-2 years approximately), and three of them deceased between 8 and 17 years due to progressive DCM. Prenatal signs were reported in a single case. The majority of them developed joint contractures after birth, as shown in Figure 2.

### Evidence on combinations of TTNtv including splice variants

Fifteen patients (group D) have bi-allelic splice variants affecting canonical exons (n=3) or a monoallelic splice variant affecting the expression of a canonical exon and a second nonsense or indel variant in canonical exons (n=12). This genotype is associated with lower lethality. There was no fetal death registered, while two of them died in the peripartum. Two cases carrying out-of-frame deletion in exon 359 in compound heterozygosity with a splice variant in a canonical exon have also been described (group F, Suppl Table 1).

The occurrence of the other signs and symptoms is summarized in Figure 2.

### Prevalence of dilated cardiomyopathy

Thirty-four cases out of 103 carried biallelic TTNtv predicted to impact both N2BA and N2B cardiac isoforms: 18 of them (53%) were reported to have cardiac involvement. In 11 cases, DCM became overt in childhood or at juvenile age, while six cases were reported to have other cardiac congenital anomalies, such as atrial septal aneurysm, atrial septal defects, large outlet and atypical muscular ventricular septal defect, left ventricular non-compaction, and left ventricular dysfunction/hypocontractility. In 16 of the 34 cases, cardiac signs or symptoms were excluded (n= 6) or not reported (n= 10).

## Discussion

The clinical spectrum of titinopathies has expanded considerably in recent years, with additional evidence of a severe end of the phenotypic spectrum caused by biallelic combinations of truncating variants. Some of these forms begin prenatally, others are recognized at birth or develop postnatally. ^3,22,12,23,24^According to the definitions reported in Human Phenotype Ontology (https://hpo.jax.org/app/), the term “congenital” should be “used for phenotypic abnormalities or diseases initially observed at the time of birth”.^25^ For abnormalities observed prior to birth (e.g., by fetal ultrasound), the term “Antenatal onset” (HP:0030674) would be more appropriate. In contrast, “infantile onset” (HP:0003593) refers to “onset of signs or symptoms of the disease between 28 days to one year of life, while “childhood onset” (HP:0011463) refers to “onset of disease at the age of between 1 and 5 years.”. Since a standardized use of terminology may facilitate the comparison between published cases as well as the understanding of the genotype-phenotype correlation, we have adopted these definitions regarding early-onset titinopathies and we encourage a standardization of the nomenclature for *TTN*-related diseases that reflects the HPO terminology.

Although it is still difficult to delineate proper genotype-phenotype correlations, our here reported antenatal cases, together with the previously reported ones, highlight some interesting recurrent clinical findings.

Fetuses with metatranscript-only variants usually display a severe phenotype, recognizable by prenatal ultrasound examination. They have the highest rate of fetal lethality and developmental anomalies, as reported in Figures 2 and 3. Three of them had anomalies also of the central nervous system reported by ultrasound examination, such as dysgenesis of the corpus callosum and abnormal liquor volume. One case who died in the peripartum presented anomalies of cerebral sulci. The severe clinical findings in the cases with the metatranscript-only variants include joint, bone and heart anomalies, such as congenital bone fractures and hypoplastic heart. These findings are coherent with the hypothesis that the metatranscript (or, probably several transcripts including a number of the so-called meta-exons) may serve as a scaffold during the sarcomere development, and that its disruption leads to severe defects in the development of the musculoskeletal system with secondary consequences for other structures. Also, in 10 cases joint hypermobility was detected after birth. When examined after birth, muscle MRI may show the typical findings of severe amyoplasia.

Curiously, some of the patients with a TTNtv in the M-band (exon 359-364) developed contractures after birth, showing a worsening trend opposite to that of patients with variants in the metatranscript, who most frequently presented antenatal arthrogryposis multiplex congenita and then might experience slight improvement.

While the clinical interpretation of metaTTNtv has been well defined in recent years, the interpretation of TTNtv in exon 359 remains challenging. Indeed, it is a very large exon, and the precise starting point of the M band has not yet been unambiguously defined. Moreover, there are only few cases reported in the literature and for most of them the clinical description is incomplete, not allowing to draw final conclusions. In general, we see that patients with one TTNtv in exon 359 in compound heterozygosity with a TTNtv in a canonical exon may have a severe antenatal or congenital onset; some of them died after delivery, while others are alive in their infancy and young age (Figure 3). While infants with one TTNtv in exon 359 may have congenital cardiac abnormalities, patients with two biallelic variants in exon 359 (n=6) have a high rate of DCM prevalence with a later onset, as half of the reported cases (n=3) died of heart failure in childhood or in their twenties, similarly to what happens in patients with biallelic TTNtv in exon 360-363.

As expected, combinations of TTNtv with at least one splice variant affecting the expression of a canonical exon (group D) were associated with less severe phenotypes. In these patients, the alleles with the splice variant produce a slightly altered transcript and protein. In general, patients carrying at least one splice variant (either affecting the expression of a meta-transcript only exon – e.g., the recurrent splice-site variant c.39974-11T>G in intron 213 – or of a canonical exon – group E) show a lower lethality ratio (Figure 3).^26^ A splice variant in compound heterozygosity with a TTNtv in a canonical exon is most probably in-frame (total absence of titin, due to two null alleles, is incompatible with embryonic development). Similarly, when two splice variants affecting a canonical exon are present, at least one of them is expected to be in-frame. However, studies on patients’ muscle tissues would be needed to determine the effects of any of these variants on splicing on the transcripts and, thereby, on the protein.

Interestingly, regarding family history, none of the parents who carry a TTNtv in a cardiac exon are reported to show clinically overt cardiomyopathy at the time of examinations. Nevertheless, in our cohort, following the recent ACMG guidelines, the proband’s parents carrying heterozygous TTNtv in exons expressed in the titin cardiac isoforms were referred to genetic counseling. In the case of family 6, we know that cardiological evaluation with echocardiography was prescribed but not yet performed; however, one of the parents, who carries a TTNtv in A-band, is reported to play sports regularly and to have been assessed as healthy by previous cardiological evaluation. It must be emphasized that in subjects in whom a truncating variant in titin is found as an incidental finding the penetrance is far from being complete, and it is reasonably lower than the penetrance reported for patients with a positive family history for cardiac diseases.^27,28^

Together with our knowledge of the titinopathies spectrum, the number of clinicians involved in the diagnosis and management of these not-so-rare diseases is constantly growing.^29,30^ Ever since TMD was discovered ^31^, it has been mainly the neurologists who dealt with titinopathies. In recent years, following the publication of the first cases of congenital titinopathies, child neurologists, neonatologists, paediatricians have been increasingly involved. The present study highlights that phenotypes that were thought to be congenital or postnatal may present typical signs as early as in the antenatal period, thus opening the possibility of early detection, higher diagnostic rate, and more in-depth investigations of the natural history of the disease. Probably, we are still missing the most severe end spectrum of titinopathies, as we are used to studying fetuses from late miscarriages, or dead infants, while only few investigations are usually performed on early miscarriages. Moreover, prenatal tests often do not include TTN sequencing.^32–34^. Titin’s involvement in prenatal and congenital phenotypes has similarities with that of severe variants in nebulin (*NEB*), a known sarcomeric protein. Recessive truncating mutations in *NEB* may cause arthrogryposis multiplex congenita, type 6 (OMIM # 619334), resulting in a clinical phenotype that is very similar to that observed in the severe cases of recessive titinopathies. Noteworthily, titin and nebulin act together as ‘molecular rulers’, respectively of the thick and the thin filaments of the sarcomere.^35^

Some recent studies focused on the ethical implications of a genetic diagnosis in severe newborns and fetal cases.^36^ In line with those reflections, although attention must be paid to offering proper genetic counseling, we believe that a prompt diagnosis can be of great benefit to the family, allowing the parents to face the future and avoid repeated negative pregnancy experiences that could have major psychological and physical repercussions.^37,38^

We are aware that our research may have some limitations. First of all, prenatal ultrasound examination is operator-dependent, and thus the prevalence of antenatal signs may be influenced by reporting bias.^39^ Also, the already published cases may be not uniformly described, as some papers were more clinically detailed than others; instead, unpublished cases were accurately detailed, but they still do not allow us to draw further genotype-phenotype conclusions. Thus, a prospective study involving many international centers would be needed to have more in-depth clinical information about severe antenatal and congenital titinopathies. Most importantly, we still have a limited knowledge of the exon usage and of the specific isoforms expressed in different prenatal and in postnatal muscles and, similarly, we lack an in-depth understanding of all the effects of truncating variants on the protein and on different transcripts.

### Conclusion

Severe recessive titinopathies, mainly caused by truncating variants in metatranscript-only exons or in exon 359, have antenatal signs resembling a syndromic phenotype, not only affecting the muscular systems but also bone, heart, and other organs, with a quite high rate of dysmorphisms. They can therefore go in differential diagnosis with several forms of arthrogryposis multiplex congenita as well as with other developmental syndromes (e.g., Noonan syndrome, Escobar syndrome, congenital myasthenic syndromes, Larsen syndrome, Pena–Shokeir syndrome or Fetal akinesia deformation sequence). It is thus crucial to raise awareness that TTNtv do not only cause myopathies, but also complex phenotypes, and that antenatal and congenital titinopathies need to be recognized in different clinical settings. As a consequence, titin should be included among the genes to be analyzed in antenatal cases with arthrogryposis, neonatal hypotonia, and the above-discussed signs and symptoms.

## Data Availability

All data produced in the present work are contained in the manuscript

## Acknowledgements

The authors thank all the patients and family members for their cooperation and all the clinicians for collecting patient data.

## Author Information

Conceptualization: M.S., M.F.D.F. Data Curation: M.F.D.F. Data Analysis: M.F.D.F., V.L. Patients Recruitment and Phenotypic Characterization: E.G., A.B., T.H., M.M., M.J., M.I., L.S., P.D., G.C.C.C, B.U. Supervision: M.S., B.U. Writing-original draft: M.F.D.F., V.L. Writing-review & editing: M.S., B.U.

## Ethics Declaration

All the patients or their legal guardians provided written informed consent to their referring clinician. The study was performed in accordance with the Declaration of Helsinki. Ethical approval was obtained through the institutional review board HUS:195/13/03/00/11 (Helsingin Yliopistollinen Sairaala - Hospital District of Helsinki and Uusimaa).

## Figure legends

Supplementary Figure 1. Pedigrees of the 10 unpublished cases. Small squares and circles indicate prenatal death.

Supplementary Table 1: Extended clinical description, TTNtv and bibliography of all published cases. “NA” means that the information was not included in the clinical description by the authors.

