## Supplementary figures and images for "The crucial role of titin in fetal development: recurrent miscarriages and bone, heart, and muscle anomalies characterize the severe end of titinopathies spectrum"

### Supplementary Figure 1

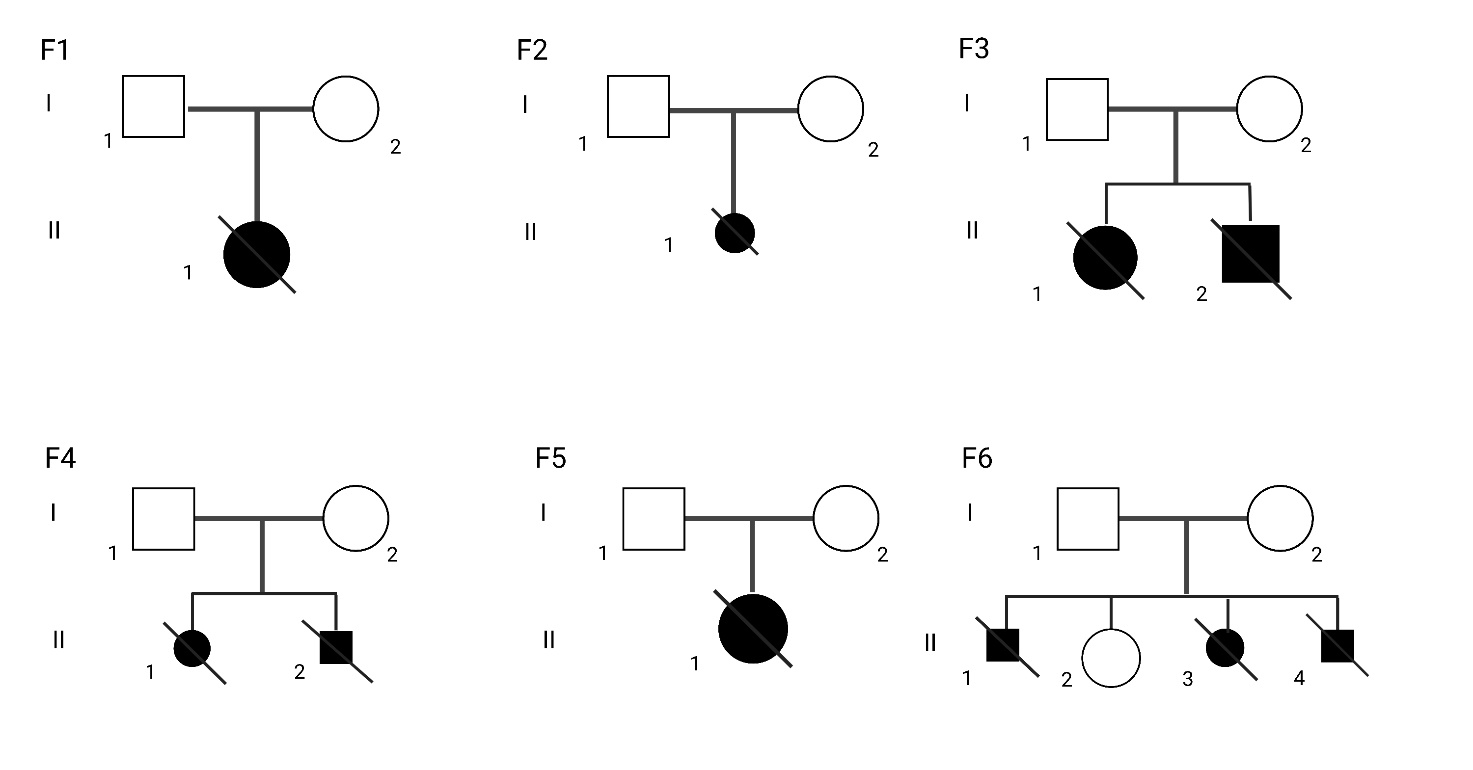

Supplementary Figure 1
